# Development of a Deep Learning Model for Early Alzheimer’s Disease Detection from Structural MRIs and External Validation on an Independent Cohort

**DOI:** 10.1101/2021.05.28.21257318

**Authors:** Sheng Liu, Arjun V. Masurkar, Henry Rusinek, Jingyun Chen, Ben Zhang, Weicheng Zhu, Carlos Fernandez-Granda, Narges Razavian, for the Alzheimer’s Disease Neuroimaging Initiative

**Affiliations:** Center for Data Science, NYU, New York, NY, USA; Center for Cognitive Neurology, Department of Neurology, NYU Grossman School of Medicine, New York, NY, USA; Neuroscience Institute, NYU Grossman School of Medicine, New York, NY, USA; Department of Radiology, NYU Grossman School of Medicine, New York, NY, USA; Department of Psychiatry, NYU Grossman School of Medicine, New York, NY, USA; Department of Population Health, NYU Grossman School of Medicine, New York, NY, USA; Courant Institute of Mathematical Sciences, NYU, New York, NY, USA

## Abstract

Early diagnosis of Alzheimer’s disease plays a pivotal role in patient care and clinical trials. In this study, we have developed a new approach based on 3D deep convolutional neural networks to accurately differentiate mild Alzheimer’s disease dementia from mild cognitive impairment and cognitively normal individuals using structural MRIs. For comparison, we have built a reference model based on the volumes and thickness of previously reported brain regions that are known to be implicated in disease progression. We validate both models on an internal held-out cohort from The Alzheimer’s Disease Neuroimaging Initiative (ADNI) and on an external independent cohort from The National Alzheimer’s Coordinating Center (NACC). The deep-learning model is more accurate and significantly faster than the volume/thickness model. The model can also be used to forecast progression: subjects with mild cognitive impairment misclassified as having mild Alzheimer’s disease dementia by the model were faster to progress to dementia over time. An analysis of the features learned by the proposed model shows that it relies on a wide range of regions associated with Alzheimer’s disease. These findings suggest that deep neural networks can automatically learn to identify imaging biomarkers that are predictive of Alzheimer’s disease, and leverage them to achieve accurate early detection of the disease.

## Introduction

Alzheimer’s disease is the leading cause of dementia, and the sixth leading cause of death in the United States ^1^. Improving early detection of Alzheimer’s disease is a critical need for optimal intervention success, as well as for counseling patients and families, clinical trial enrollment, and determining which patients would benefit from future disease-modifying therapy ^2^. Alzheimer’s disease related brain degeneration begins years before the clinical onset of symptoms. In recent years, the development of PET imaging techniques using tracers for amyloid and tau have improved our ability to detect Alzheimer’s disease at preclinical and prodromal stages, but they have a significant disadvantage of being expensive and requiring specialized tracers and equipment. Many studies have shown that structural MRI-based volume measurements, particularly of the hippocampus and medial temporal lobe, are somewhat predictive of Alzheimer’s disease progression ^3–7 8–12,13^. While the availability and cost of MRI is beneficial, these early attempts to discriminate healthy aging from Alzheimer’s disease based on volumetry had significant limitations, including small sample size and reliance on semi-automated segmentation methods. This motivated the emergence of more sophisticated methods to analyze MRI data based on machine learning.

In the last decade, machine learning and fully automatic segmentation methods have achieved impressive results in multiple computer vision and image processing tasks. Early applications of machine learning to Alzheimer’s disease diagnosis from MRIs were based on discriminative features selected a priori ^14–17^. These features include regional volumes and cortical thickness segmented from brain regions known to be involved/implicated with memory loss and accelerated neurodegeneration that accompany Alzheimer’s disease ^17,18,19^. Newer machine learning methods based on deep convolutional neural networks (CNNs) make it possible to extract features directly from image data in a data-driven fashion. These methods have been shown to outperform traditional techniques based on predefined features in most image processing and computer vision tasks ^20,21^. In the biomedical field, CNN-based methods also have the potential to reveal new imaging biomarkers ^22,23^. Multiple studies have addressed mild Alzheimer’s disease dementia detection from MRI via deep learning, with notable examples of 3D convolutional neural networks based on 3D AlexNet, 3D Resnet, patch based models, Siamese networks, auto-encoder based models, among others ^24–26^. Based on systematic reviews and survey studies ^27,28^, many of previous approaches had major limitations in their design or validation: Most of these studies focus on distinguishing Alzheimer’s disease dementia patients from normal controls. However, in order to develop effective and clinically relevant early detection methods, it is crucial to also differentiate prodromal Alzheimer’s disease, otherwise known as mild cognitive impairment (MCI), from both normal controls and patients with manifest Alzheimer’s disease dementia. Some recent studies have made inroads to this end ^29–31^, but do not evaluate their results on large independent cohorts where there can be more variability in image acquisition and clinical diagnosis, more representative of real world scenarios. The goal of this work is to address these significant challenges.

We propose a deep-learning model based on a novel CNN architecture that is capable of distinguishing between persons who have normal cognition, MCI, and mild Alzheimer’s disease dementia. The proposed model is trained using a publicly available dataset from the Alzheimer’s Disease Neuroimaging Initiative (ADNI). Although a multisite study, ADNI sites follow a rigorous standard protocol and stringent quality control to minimize site differences and improve our ability to reliably detect neuroanatomical changes. To assess the performance of the proposed methodology when applied in more realistic conditions, we evaluated our approach on an entirely independent cohort of 1522 subjects from the National Alzheimer’s Coordinating Center (NACC). Since (until very recently) each NIH/NIA funded center contributing to the NACC database is free to employ different acquisition parameters, this enables us to validate our approach on imaging data acquired with variable and non standardized protocols.

Our approach achieves an area-under-the-curve (AUC) of 85.12 (95% CI: 84.98 - 85.26) when distinguishing between cognitive normal subjects and subjects with either MCI or mild Alzheimer’s dementia in the independent NACC cohort. For comparison, we have built a reference model based on the volumes and thickness of previously reported brain regions that are known to be implicated early in disease progression. These measures were obtained by the automated segmentation tool Freesurfer ^32^. We demonstrate that our proposed deep-learning model is more accurate and orders-of-magnitude faster than the ROI-volume/thickness model. Our results suggest that CNN-based models hold significant promise as a tool for automatic early diagnosis of Alzheimer’s disease across multiple stages.

## Results

### Study participants

The study is based on data from ADNI and NACC. The cohorts are described in Table 1. ADNI is a longitudinal multicenter study designed to develop clinical, imaging, genetic, and biochemical biomarkers for the early detection and tracking of Alzheimer’s disease ^33^. NACC, established in 1999, is a large relational database of standardized clinical and neuropathological research data collected from Alzheimer’s disease centers across the USA ^34^. Both datasets contain MRIs labeled with one of three possible diagnoses based on the cognitive status evaluated closest to the scanning time: cognitive normal (CN), mild cognitive impairment (MCI), or Alzheimer’s disease dementia. Labeling criteria are included/described in supplementary table S1.

**Table 1:**
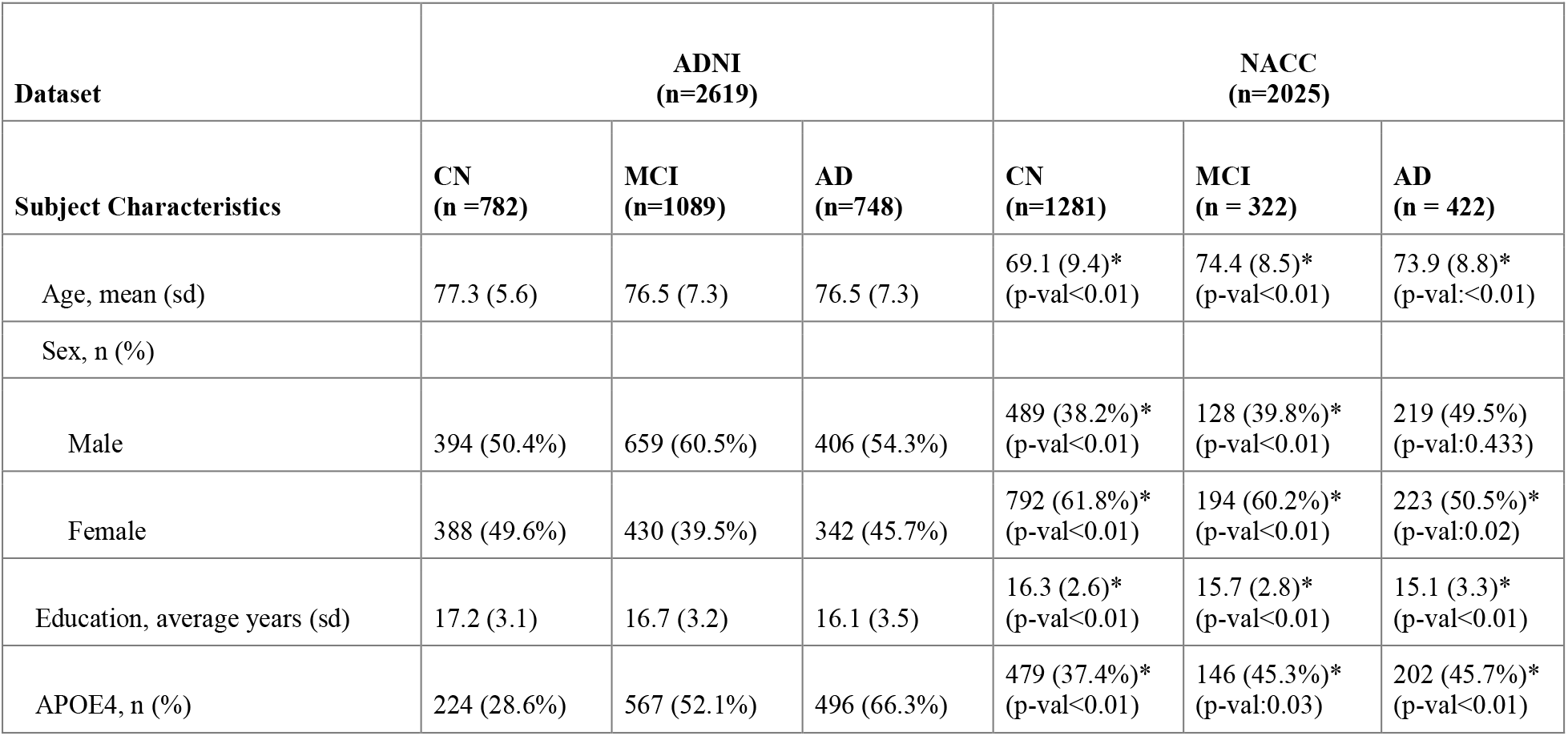
Demographic characteristics of the ADNI and NACC cohorts. Significance of differences between NACC and ADNI for each cognitive category are reported. Statistical significance (at p-value < 0.05) is indicated by *.

We separated the ADNI subjects at random into three disjoint sets: a training set, with 1939 scans from 463 individuals, a validation set with 383 scans from 99 individuals, and a test cohort of 297 scans from 90 individuals. We built an additional independent test cohort based on NACC using the following inclusion criteria: individuals aged ≽ 55 years with MRIs within ± 6 months from the date of clinically-confirmed diagnosis of cognitively normal (CN), mild cognitive impairment (MCI), or mild Alzheimer’s disease dementia (AD). This resulted in a cohort of 1522 individuals (1281 CN, 322 MCI and 422 AD) and 2045 MRIs.

Table 1 reports the basic demographic and genetic characteristics of participants whose scans were used in this study. While cognitive groups in ADNI are well matched on age, in NACC cohort CN subjects were on the average ∼ 5 years younger than the two impaired groups; they were 6-7 years younger than ADNI participants. There is a female predominance in NACC data, especially in CN and MCI, and a male predominance in ADNI, notable in the impaired stages. In the impaired population (MCI, AD), the prevalence of the AD genetic risk factor APOE4 is lower in NACC compared to ADNI. Considering these significant differences in cohort characteristics, using NACC as an external validation cohort allows us to assess the robustness of our method. In both ADNI and NACC, education seems to also be lower with progressive impairment stage, which may be indicative of lower structural reserve.

### Models

Our deep-learning model is a 3D convolutional neural network (CNN), with an architecture that is specifically optimized for the task of distinguishing CN, MCI, and AD status based on MRIs (Figure 1.b., see the Methods section for more details). We also designed a gradient-boosting model^35^ based on 138 volumes and thickness of clinically-relevant brain ROIs (see supplementary Table S2 for the list) obtained by segmenting the MRIs using the Freesurfer software (v6.0, surfer.nmr.mgh.harvard.edu). Quality control was applied to the segmentations through sampling and visual inspection by a trained neuroimaging analyst (JC), in consultation with a clinical neurologist (AVM). Details of the quality control process are included in the Methods section.

**Figure 1:**
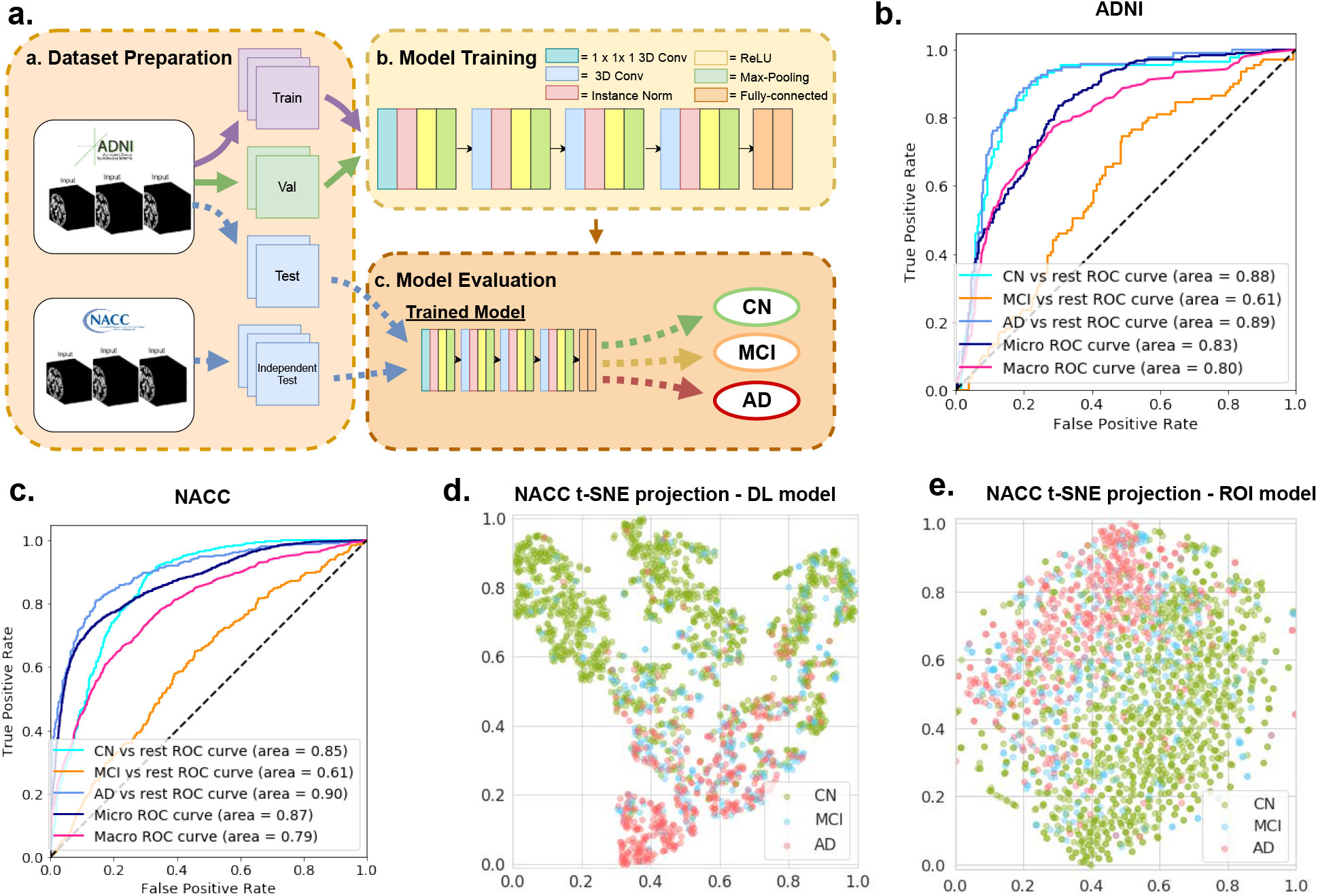
Overview of the deep learning framework and performance for Alzheimer’s automatic diagnosis. (**a**) Deep learning framework used for automatic diagnosis. (**b**) Receiver operating characteristic (ROC) curves for classification of cognitively normal (CN), mild cognitive impairment (MCI) and Alzheimer’s disease (AD), computed on the ADNI held-out test set. (**c**) ROC curves for classification of cognitively normal (CN), mild cognitive impairment (MCI) and Alzheimer’s disease (AD) on the NACC test set. (**d**) Visualization using t-SNE projections of the features computed by the proposed deep-learning model. Each point represents a scan. Green, blue, red colors indicate predicted cognitive groups. CN and AD scans are clearly clustered. (**e**) Visualization using t-SNE projections of the 138 volumes and thickness in the ROI-volume/thickness model. Compared to (d) the separation between CN and AD scans is less marked. The t-SNE approach is described in details in the methods section.

### Identification of cognitive impairment status

In order to evaluate the diagnostic performance of the machine-learning models, we computed ROC curves for the task of distinguishing each class (CN, MCI, or AD) from the rest. Table 2 includes our results. On the ADNI held-out test set, the proposed deep-learning model achieved the following AUCs: 87.59 (95% CI: 87.13 - 88.05) for CN vs the rest, 62.59 (95% CI: 62.01 - 63.17) for MCI vs the rest, and 89.21 (95% CI: 88.88 - 89.54) for AD vs the rest.

**Table 2:** Classification performance in ADNI held-out set and an external validation set. Area under ROC curve for classification performance based on the deep learning model vs the ROI-volume/thickness model, for ADNI held-out set and NACC external validation set. Deep learning model outperforms ROI-volume/thickness-based model in all classes.

|  | ADNI held-out<br>(n=90 individuals, 297 scans) |  | NACC external validation<br>(n=1522 individuals, 2025 scans ) |  |
| --- | --- | --- | --- | --- |
|  | Deep learning model<br>Area under ROC curve | ROI-volume/thickness<br>Area under ROC curve | Deep learning model<br>Area under ROC curve | ROI-volume/thickness<br>Area under ROC curve |
| Cognitively Normal | 87.59<br>(95% CI: 87.13 - 88.05) | 84.45<br>(95% CI: 84.19 - 84.71) | 85.12<br>(95% CI: 85.26 - 84.98) | 80.77<br>(95% CI: 80.55 - 80.99) |
| Mild Cognitive<br>Impairment | 62.59<br>(95% CI: 62.01 - 63.17) | 56.95<br>(95% CI: 56.27 - 57.63) | 62.45<br>(95% CI: 62.82 - 62.08) | 57.88<br>(95% CI: 57.53 - 58.23) |
| Alzheimer’s Disease<br>Dementia | 89.21<br>(95% CI: 88.88 - 89.54) | 85.57<br>(95% CI: 85.16 - 85.98) | 89.21<br>(95% CI: 88.99 - 89.43) | 81.03<br>(95% CI: 80.84 - 81.21) |

The AUCs of the ROI-volume/thickness model were statistically significantly lower than those of the deep-learning model: 84.45 (95% CI: 84.19 - 84.71) for CN vs the rest, 56.95 (95% CI: 56.27 - 57.63) for MCI vs the rest, and 85.57 (95% CI: 85.16 - 85.98) vs the rest.

The deep-learning model achieved similar performance on the NACC external validation data compared to ADNI held-out test set, achieving AUCs of 85.12 (95% CI: 85.26 - 84.98) for CN vs the rest, 62.45 (95% CI: 62.82 - 62.08) for MCI vs the rest, and 89.21 (95% CI: 88.99 - 89.43) for AD vs the rest. The ROI-volume/thickness model suffered a more marked decrease in performance, achieving AUCs of 80.77 (95% CI: 80.55 - 80.99) for CN vs the rest, 57.88 (95% CI: 57.53 - 58.23) for MCI vs the rest, and 81.03 (95% CI: 80.84 - 81.21) for AD vs the rest. In Supplementary Figure F1 we also report the precision-recall curve for the deep learning model on both the ADNI held-out test set and NACC external validation set. In Supplementary Table S5 and S6 we provide confusion matrices that show the misclassification rate between different classes.

We analyze the features extracted by the deep-learning model using t-distributed stochastic neighbour embedding (t-SNE), which is a projection method suitable for data with high-dimensional features, such as those learned via deep learning^36^. Figure 1(d) shows the two dimensional t-SNE projections of the deep learning based features corresponding to all subjects in the NACC dataset. Points corresponding to CN and AD subjects are well separated. Figure 1(e) shows the t-SNE projections of the ROI-volume/thickness features. In this case the separation between AD and CN scans is less clear. In both cases, points corresponding to MCI scans are not clustered together. This visualization is consistent with our results, which suggest that the features extracted by the deep-learning model are more discriminative than the ROI-based features, and also that distinguishing individuals diagnosed as MCI is more challenging.

Our deep learning model is significantly faster than classification based on regions of interest. On average, for each MRI, our deep learning model requires 0.07s (plus 7 min for NMI normalization as preprocessing), compared to 11.2 hours required for extracting the regions of interest with Freesurfer.

### Progression Analysis

We investigated whether the deep-learning model and ROI-volume/thickness model learn features that may be predictive of cognitive decline of MCI subjects to AD. In the held-out test set of ADNI, we divided the subjects into two groups based on the classification results of the deep learning model from the baseline date defined as the time of initial diagnosis as MCI: group A if the model classified the first scan as AD (n=18), and group B if it did not (n=26). Figure 2 shows the proportion of subjects in each group who progressed to AD at different months past the baseline. Based on the deep learning model, 23.02% (95% CI: 21.43% - 24.62%) of subjects in group A (blue line) progress to AD, compared to 8.81% (95% CI: 8.09% - 9.53%) of subjects in group B (red line). For the ROI-volume/thickness model, 20.22% (95% CI: 18.49% - 21.95%) of subjects in group A (blue line) progress to AD, compared to 11.11% (95% CI: 10.32% - 11.89%) of subjects in group B (red line). The forecasting ability of the deep learning model is therefore significantly higher than that of the ROI-volume/thickness based model. Our results suggest that deep-learning models could be effective in forecasting the progression of Alzheimer’s disease.

**Figure 2:**
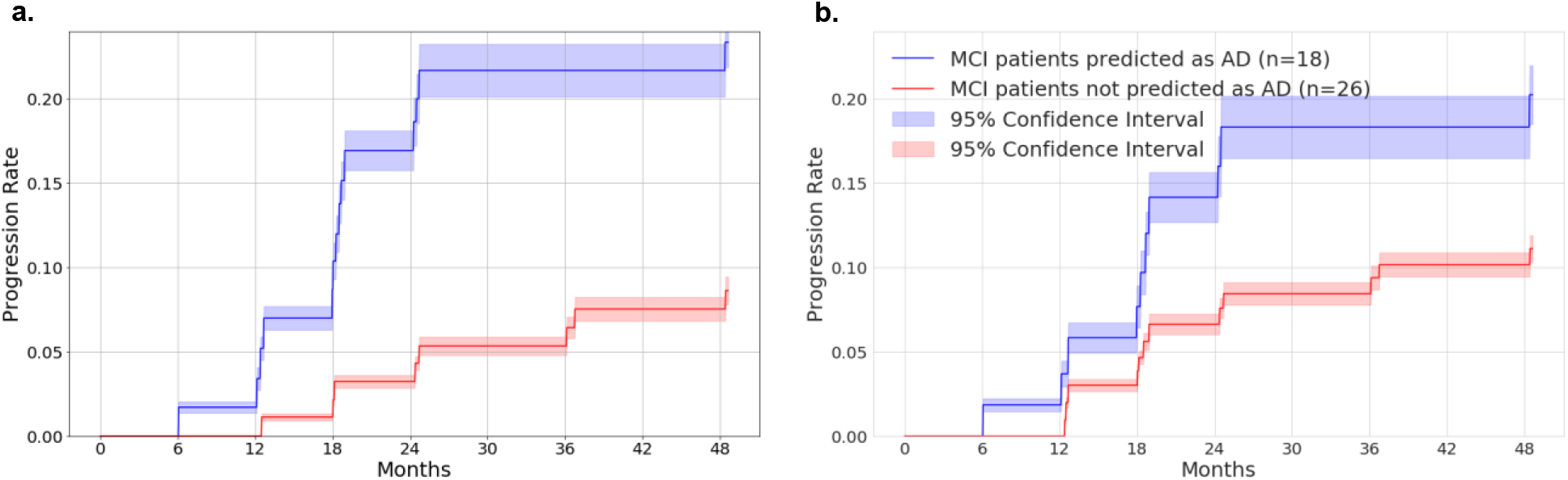
Progression analysis for MCI subjects. a) Progression analysis based on the deep learning model. b) Progression analysis based on the ROI-volume/thickness model. The subjects in the ADNI test set are divided into two groups based on the classification results of the deep learning model from their first scan diagnosed as MCI: group A if the prediction is AD, and group B if it is not. The graph shows the fraction of subjects that progressed to AD at different months following the first scan diagnosed as MCI for both groups. Subjects in group A progress to AD at a significantly faster rate, suggesting that the features extracted by the deep-learning model may be predictive of the transition.

### Sensitivity to Group Differences

Figure 3 shows the performance of the deep learning and the ROI-volume/thickness model across a range of sub-cohorts based on age, sex, education and APOE4 status. Supplementary table S3 includes the AUC values and 95% confidence intervals. The deep learning model achieves statistically significantly better performance on both ADNI and NACC cohorts. One exception is the ApoE4-positive group within NACC, for which classification of CN vs the rest, and MCI vs the rest by deep learning were worse than the ROI-volume/thickness model. Differences in sex and age representation in NACC versus ADNI, as discussed above, could influence this result. However, deep learning outperformed the ROI-volume/thickness model in both males and females, in both cohorts. The CN cohort in the NACC dataset is on average younger than that in the ADNI dataset (Table 1). In order to control for the influence of age, we stratified the NACC cohort into two groups (above and below the median age of 70 years old). However, the AUCs of the deep learning model for classification of CN vs the rest were very similar in both groups (85.2 for the younger cohort, to 86.1 for the older cohort). Another possible explanation for the ApoE4 difference is that NACC has a more clinically heterogeneous population, including participants with early stages of other diseases for which ApoE4 can be a risk factor, such as Lewy body and vascular dementia, and which can be clinically indistinguishable from AD at CN and MCI stages. In both ADNI and NACC, low education (< 15 years) also identified a subgroup in which deep learning was outperformed by ROI-volume/thickness model. This subgroup’s clinical presentation, especially at prodromal stages, may be more directly related to brain volume changes according to the cognitive reserve hypothesis ^37^.

**Figure 3:**
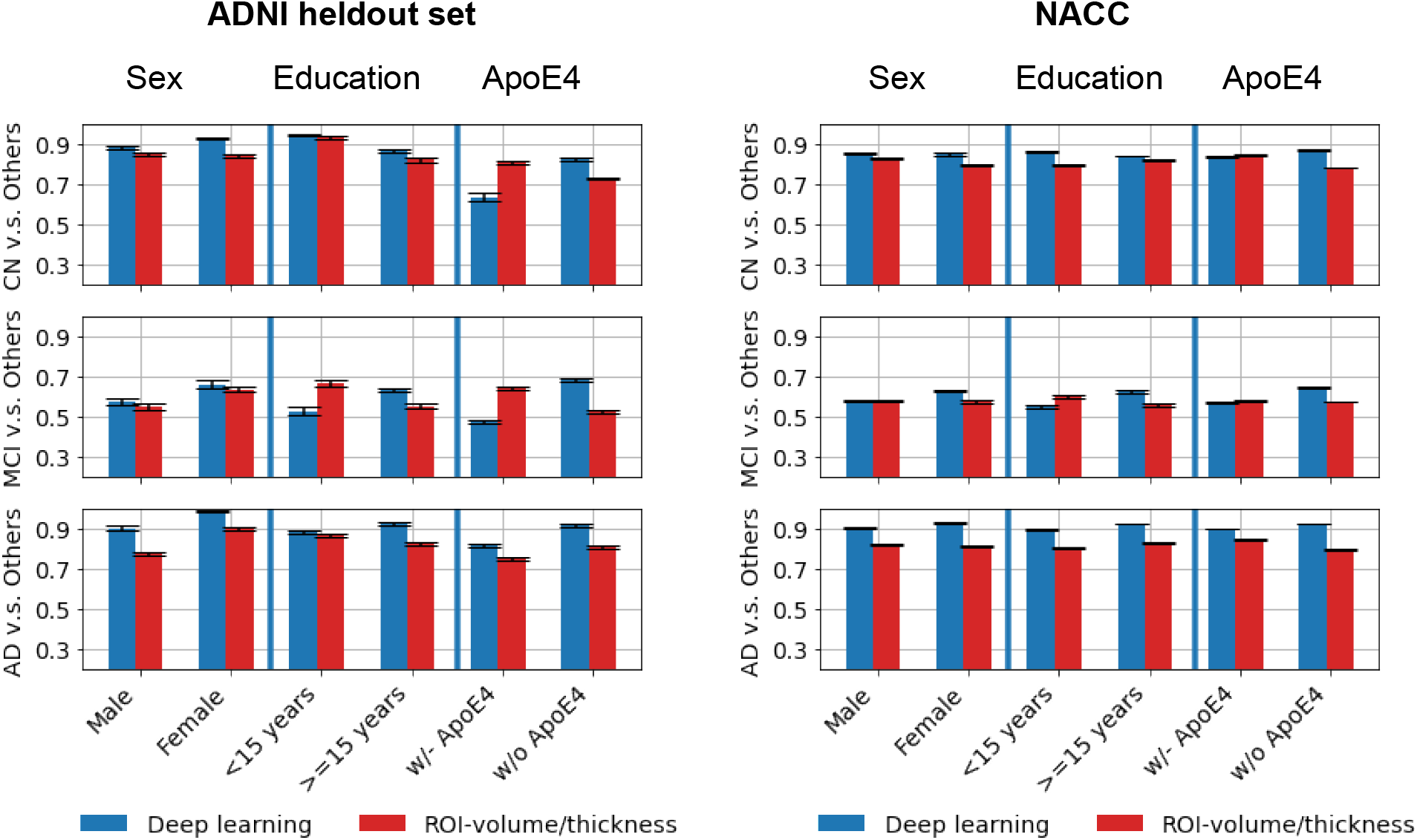
Performance across different subgroups. Performance of the deep learning model (in blue) and of the ROI-volume/thickness model (in red) for different subpopulations of individuals, separated according to sex, education, and ApoE4 status.

### Impact of dataset size

In order to evaluate the impact of the training dataset size, we trained the proposed deep-learning model and the ROI-volume/thickness model on datasets of varying sizes, by randomly subsampling the training dataset. As shown in Figure 4, the performance of the ROI-volume/thickness model improves when the training data increases from 50% to 70%, but remains essentially stagnant after further increases. In contrast, the performance of the deep learning model consistently improves as the size of the training set increases.

**Figure 4:**
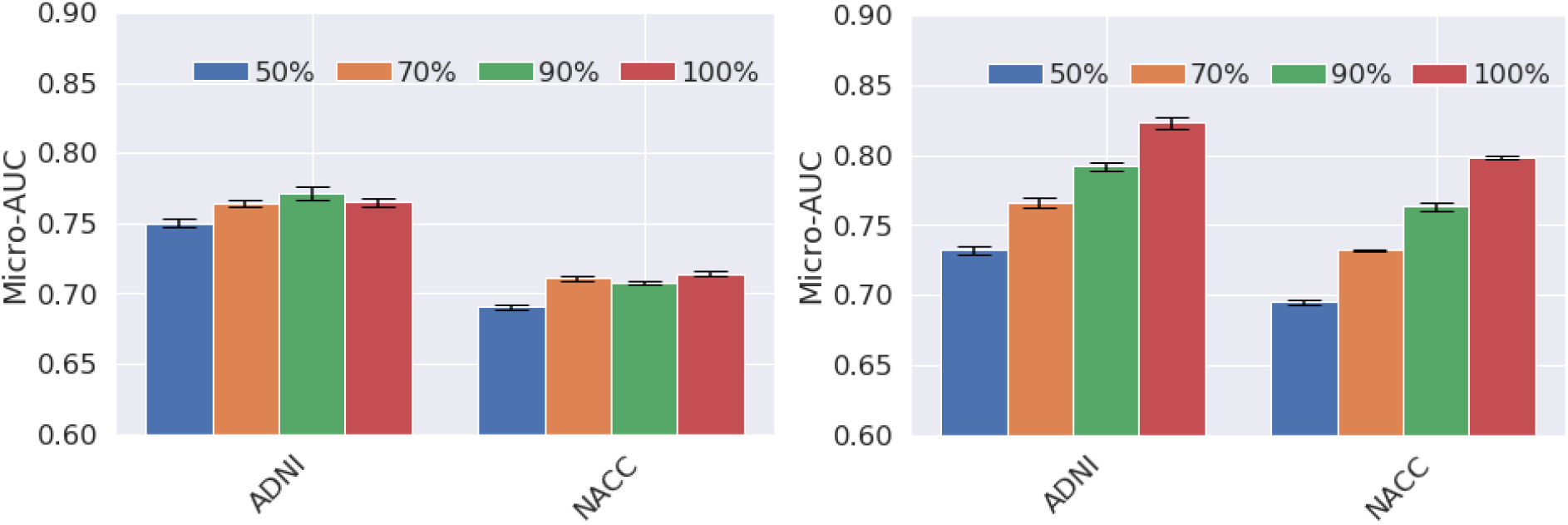
Datasize Impact. Performance of the baseline ROI-volume/thickness model (left) and the proposed deep learning model (right) when trained on datasets with different sizes (obtained by randomly subsampling the training set). The performance of the ROI-volume/thickness model improves when the training data increases from 50% to 70%, but remains essentially stagnant after further increases. In contrast, the performance of the deep learning model consistently improves as the size of the training set increases. Given that the deep learning model is trained on a very small dataset compared to standard computer-vision tasks for natural images, this suggests that building larger training sets is a promising avenue to further improving performance.

### Model interpretation

In order to visualize the features learned by the deep learning model, we computed saliency maps highlighting the regions of the input MRI scans that most influence the probability assigned by the model to each of the three classes (CN, MCI, or AD), as described in the Methods section. Figure 5 shows the saliency maps corresponding to each class, aggregated over all scans in the ADNI held-out test set. The figure also reports the relative importance of the top 30 ROIs, quantified by a normalized count of voxels with high gradient magnitudes (see Methods section for more details). In Supplementary Table S4 we report the full list, as well as a quantification of the importance of the ROIs for the baseline volume/thickness model.

**Figure 5:**
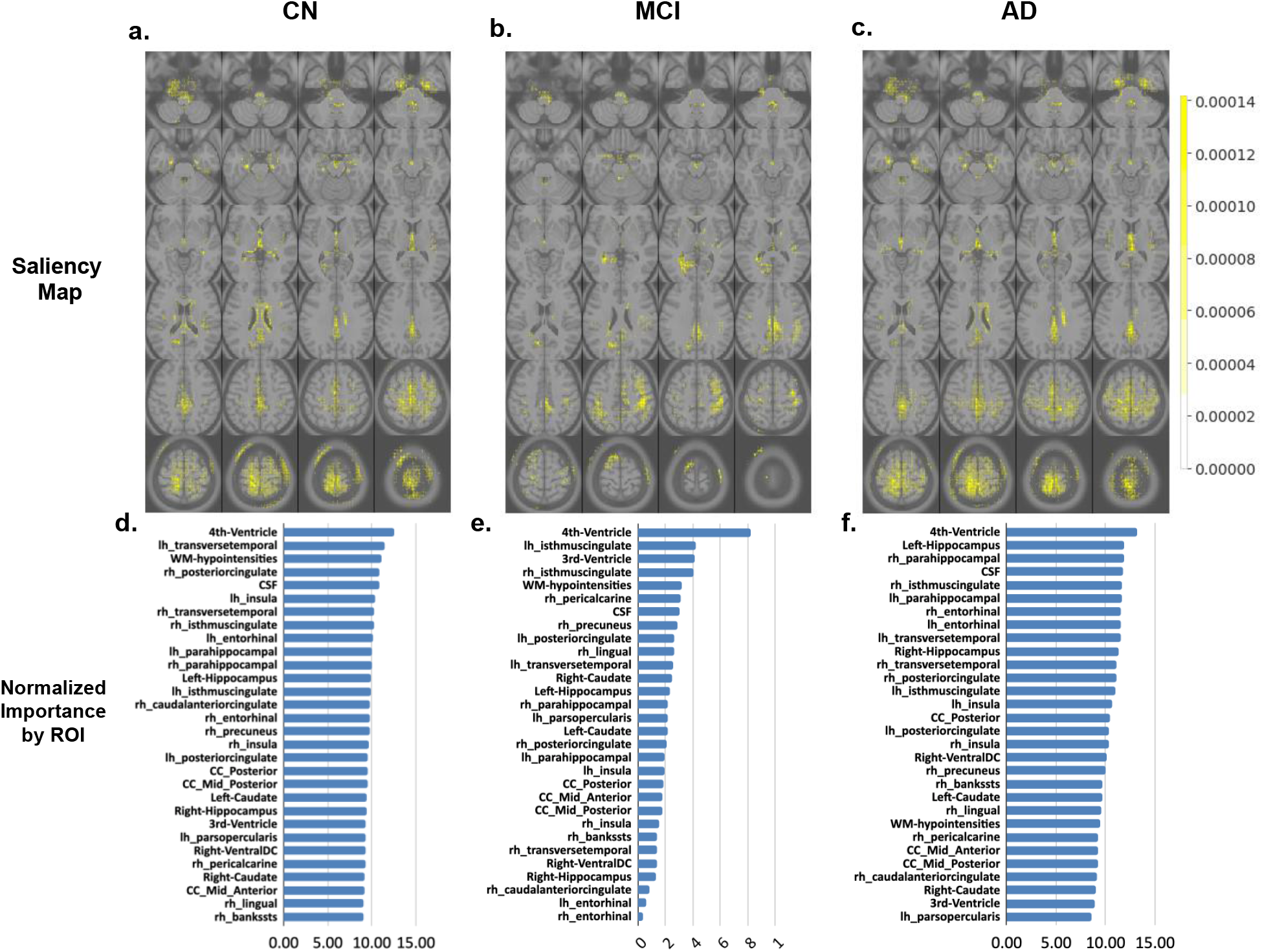
(a-c) Visualization of the aggregated importance of each voxel (in yellow) in the deep learning model when classifying subjects into CN/MCI/AD. For each subject, the importance map was computed using the gradient of the deep-learning model with respect to its input (Details in Methods section). The computed gradients are visualized over the MNI T1-weighted template. (d-f) Top 30 regions of interest sorted by their normalized gradient count, which quantifies their importance (see Methods section), for each of the classes.

### Combining deep learning and the ROI-volume/thickness models

We combined the deep learning and ROI-volume/thickness model model by treating the predictions of the deep learning model as new features, and fusing them with the volume/thickness features to train a new gradient boosting model. This model achieved the following AUCs on ADNI test set: 89.25 (95% CI: 88.82 - 89.63) for CN vs the rest, 70.04 (95% CI: 69.40 - 70.68) for MCI vs the rest, and 90.12 (95% CI: 89.75 - 90.49) for AD vs the rest. It achieved similar performance on the NACC external validation data: AUCs of 85.49 (95% CI: 85.06 - 85.92) for CN vs the rest, 65.85 (95% CI: 65.37 - 66.33) for MCI vs the rest, and 90.12 (95% CI: 89.86 - 90.38) for AD vs the rest.

## Discussion

Our analysis supports the feasibility of automated MRI-based prioritization of elderly patients for further neurocognitive screening using deep learning models. Recent literature ^38^ has consistently shown high prevalence of missed and delayed diagnosis of dementia in primary care. Major contributory factors include insufficient training, lack of resources, and limited time to comprehensively perform early dementia detection. We show that deep learning is a promising tool to perform automatic early detection of Alzheimer’s disease from MRI data. The proposed model is able to effectively identify CN and AD subjects based on MRI data, clearly outperforming the model based on more traditional features such as ROI volumes and thicknesses. While identifying MCI is more challenging, our method still demonstrates improved performance compared to traditional methods. Moreover, as demonstrated in Figure 3, MCI misclassification as mild Alzheimer’s dementia may prove to be clinically useful in identifying a higher risk MCI subgroup that progresses faster. For example, in practice, if subsequent functional assessment confirms MCI rather than dementia, these patients may need to be monitored more closely, counseled differently, and more quickly introduced to disease-modifying therapy that may be available in the future..

Our results suggest that deep convolutional neural networks automatically extract features associated with Alzheimer’s disease. The analysis of feature importance for the ROI-volume/thickness method shows that the importance of the left hippocampus is an order of magnitude larger than any of the other ROIs, suggesting that this region may dominate the output of the model (see Supplementary Table S4). In contrast, the deep-learning model exploits a much wider range of regions. This highlights the potential of such models to exploit features in imaging data, which are not restricted to traditional measures such as volume and thickness. Many regions previously implicated in distinguishing stage severity of Alzheimer’s disease are recognized as salient by the deep-learning model (see Results). The left and right entorhinal cortex and hippocampus, which are considered the most relevant ROIs to the early stages of Alzheimer’s disease progression by the Braak staging method^39^, appear within the 11 most salient regions. Other regions identified as salient that are in agreement with previous literature include the inferior lateral ventricles (left and right), parahippocampal gyrus (left and right), and white matter hypo-intensities ^40^. When comparing to another study that used segmented volumes to distinguish Alzheimer’s disease dementia from controls ^41^, the 4th ventricle was uniquely and highly relevant to the deep-learning model whereas certain gyri (fusiform, temporal, angular, supramarginal) were not identified as particularly salient.

Our results suggest several avenues for improving deep learning models for early detection of Alzheimer’s disease. First, the available datasets to train these models is quite limited compared to standard benchmarks for computer vision tasks, which have millions of examples ^42^. We show that the number of training data has a strong effect on performance, so gathering larger training sets is likely to produce a significant boost in accuracy. Second, we have shown that combining features learned using deep learning with more traditional ROI-based features such as volume and thickness improves the performance. However, using segmentation-based features is very costly computationally (segmentation takes 11.2 hours on average, compared to the 7.8 minutes needed to apply the deep-learning model). Designing deep-learning models trained to extract volumetric information automatically may improve performance, without incurring such a heavy computational cost.

In this work, we limit our analysis to brain structural MRIs, in order to develop imaging biomarkers for early detection of Alzheimer’s disease. Integrating information such as age or education, genetic data (e.g. single nucleotide polymorphisms), clinical test data from electronic health records, and cognitive performance tests results could provide a more holistic view of Alzheimer’s disease staging analysis. Building deep learning models capable of effectively combining such information with imaging data is an important direction for future research.

## Methods

### Data

The data used in this study consists of imaging and diagnosis data from Alzheimer’s Disease Neuroimaging Initiative (ADNI) and National Alzheimer’s Coordinating Center (NACC). The structural MRI scans (T1 MRIs) were downloaded from the ADNI portal^2^ (n=2619). As the diagnoses are done for each screen visit, we directly used the current diagnosis (DXCURREN column), in the ADNI’s diagnosis summary table for each scan.

We used the NACC dataset for external evaluation (n=2025 MRI scans). The NACC initiative was established in 1999, and maintains a large relational database of standardized clinical and neuropathological research data collected from Alzheimer’s disease centres across the USA ^43^. The scan-level diagnostic labels were obtained based on diagnosis within 6 months of the scanning time (closest visit). Scans which did not have any diagnostic information within 6 month of the scan were excluded. Volumetric data for the same cohort were compiled from Freesurfer outputs (with built-in commands asegstats2table and aparcstats2table).

Our analysis was restricted to patients over the age of 55 in both cohorts, and we only considered T1 MRIs (without contrast) for the study.

### MRI Data preprocessing

All scans in both cohorts were preprocessed by applying bias correction and spatial normalization to the Montreal Neurological Institute (MNI) template using the Unified Segmentation procedure ^44^ as implemented in step A of the T1-volume pipeline in the Clinica software ^45^. The preprocessed images consist of 121 × 145 × 121 voxels, with a voxel size of 1.5 × 1.5 × 1.5 *mm*^*3*^.

### Deep learning model

A 3D CNN, composed of convolutional layers, instance normalization ^46^, ReLUs and max-pooling layers, was designed to perform classification of Alzheimer’s disease and mild cognitive impairment and normal cognition cases. The architecture is described in more detail in Figure 1.a.(b). In a preliminary work we showed that the proposed architecture is superior to state-of-the-art CNNs for image classification^31^. The proposed architecture contains several design choices that are different from the standard convolutional neural networks for classification of natural images: 1) instance normalization, an alternative to batch normalization ^47^ introduced originally in the context of style transfer; 2) small kernel and stride in the initial layer; 3) wider network architecture with more filters and less layers. These techniques all independently contribute to boost performance.

As is standard in deep learning for image classification ^48^, we performed data augmentation via Gaussian blurring with mean zero and standard deviation randomly chosen between 0 and 1.5, and via random cropping (using patches of size 96 × 96 × 96).

Training and testing routines for the DL architectures were implemented on an NVIDIA CUDA parallel computing platform (accessing 2 Intel(R) Xeon(R) Gold 6230 CPU @ 2.10 GHz nodes on the IBM LSF cluster each with 2 NVIDIA Tesla V100 SXM2 32 GB GPUs) using GPU accelerated NVIDIA CUDA toolkit (cudatoolkit)^3^, CUDA Deep Neural Network (cudnn) and PyTorch ^49^ tensor libraries. The model was trained using stochastic gradient descent with momentum 0.9 (as implemented in the torch.optim package) to minimize a cross-entropy loss function. We used a batch size of 4 and a learning rate of 0.01 with a total 60 epochs of training. During training, the model with the lowest validation loss was selected.

### ROI-volume/thickness model

To build a model based on traditionally and commonly used ROI thickness and volumes, we first segmented each brain MRI using Freesurfer, and then computed volume and thickness from these ROIs (using the Freesurfer commands **asegstats2table** and **aparcstats2table**). In order to get the volumetric data for each scan, we processed ADNI and NACC datasets with “recon-all” command at a high performance computer cluster. 16 parallel batch jobs were carried out together, each job was assigned with 320 RAM, 40 CPUs. The average processing time for each scan is about 12 hours. 2730 scans of ADNI and 2999 of NACC were successfully processed.

For each brain MRI, a total of 138 MRI volume and thickness features (full list in Supplementary Table S2), were used as inputs to construct a Gradient Boosting (GB) classifier to predict Alzheimer’s disease statuses. GB is a standard method to leverage pre-selected features as opposed to learning them from the data ^50^. It constructs an ensemble of weak predictors by iteratively combining weaker base predictors in a greedy manner. We applied the implementation in the Python Sklearn package v0.24.1 *sklearn*.*ensemble*.*GradientBoostingClassifier* ^*51*^. We set the learning rate to 0.1 (this value was selected based on validation performance). Other hyperparameters were set to their default values. After hyperparameter selection, we trained the model 5 times with different random seeds and reported average performances of these 5 models on the ADNI test set and the external NACC test dataset.

### Quality control for Freesurfer segmentation

Because of the large number of scans, we developed a two-stage approach for the quality control (QC) of a specific ROI. In the first stage, we located outlier cases within each cohort by fitting a Gaussian distribution to the volumes and centroids of all the segmented ROIs, using a cut-off of mean +/-3 standard deviations. In the second stage, we conducted QC on the outlier and non-outlier cases separately. For the outliers, all cases were examined visually. For the non-outliers, 100 cases were randomly selected for visual examination. The visual examination was conducted by a trained neuroimaging researcher (JC), in consultation with a neurologist (AM) and a radiologist (HR). This two-stage approach was then repeated for two representative ROIs, namely hippocampus and entorhinal cortex, on each hemisphere, for each cohort. As a result, several segmentation errors were found in the outlier group of both ADNI and NACC cohorts (and excluded from follow-up machine-learning analyses), while no errors were found in the non-outlier group.

### Performance metrics

We computed areas under the ROC curve (AUC), which are widely used for measuring the predictive accuracy of binary classification problems. This metric indicates the relationship between the true positive rate and false positive rate when the classification threshold varies. As AUC can only be computed for binary classification, we computed AUCs for all three binary problems of distinguishing between one of the categories and the rest. We also calculated two types of averages, micro- and macro-average denoted as Micro-AUC and Macro-AUC respectively. The micro average treats the entire set of data as an aggregated results, and consists of the sum of true positive rates divided by the sum of false positive rates. The macro average is computed by calculating the AUC for each of the binary cases, and then averaging the results.

### t-SNE Projection

t-SNE is a tool to visualize high-dimensional data. It converts similarities between data points to joint probabilities and minimizes the Kullback-Leibler divergence between the joint probabilities of the low-dimensional embedding and the high-dimensional data. We applied the implementation in the Python Sklearn package v0.24.1 *sklearn*.*manifold*.*TSNE* with default hyperparameters.

### Interpretation of models in terms of ROIs

In order to analyze the features learned by the deep learning model, we computed saliency maps consisting of the magnitude of the gradient of the probability assigned by the model to each of the three classes (CN, MCI, or AD) with respect to its input ^52^. Intuitively, changes in the voxel intensity of regions where this gradient is large have greater influence on the output of the model. As mentioned above, we segmented the MRI scans in our dataset to locate ROIs. To determine the relative importance of these regions for the deep-learning model, we calculated the total count of voxels where the gradient magnitude is above a certain threshold (*10*^−*3*^, which is the magnitude observed in background regions where no brain tissue is present) within each ROI, and normalized it by the total number of voxels in the ROI. We excluded left-vessel, right-vessel, optic-chiasm, left-inf-lat-vent, and right-inf-lat-vent, due to their small size (less than 120 voxels).

For the ROI-volume/thickness models, we determined feature importance using a standard measure for gradient boosting methods^53^. This is obtained from the property *feature_importances_* in the Python Sklearn package v0.24.1 *sklearn*.*ensemble*.*GradientBoostingClassifier*.

### Statistical Comparisons

To report statistical significance of descriptive statistics we employed 2-tailed, unpaired testing. We used python statsmodel v0.12.2 and scipy.stats v1.6.1. A p-value < 0.05 was reported as significant. To compute 95% confidence intervals, the bootstrapping method with 100 bootstrap iterations was used.

### Reproducibility

The trained deep learning model, and corresponding code, notebooks, the IDs of subject-splits (training/validation/held-out) from publicly available ADNI, and the IDs of NACC participants included in our external validation study are all publicly available in our open-source repository: https://github.com/NYUMedML/DeepDementia.

## Data Availability

Both cohorts used in our study are publicly available at no cost upon completion of the respective data-use agreements. We used all the T1 MRI scan and clinical data from ADNI (June 15th 2019 freeze), and NACC (Jan 5th 2019 freeze). The IDs of patients and scans used in our study and training, validation, test indicators are available in our github repository. Results of our freesurfer segmentation (results of over 60,000 compute time on our HPC) for all of NACC and ADNI scans are also publicly available at no cost through NACC and ADNI websites, to anyone who has signed ADNI and NACC data-use agreements. Our trained model, training and validation scripts, and model predictions for each scan ID and patient ID, as well as freesurfer segmentation volume and thickness features are also available as open-source on github.

## Ethics

The datasets used in this analysis are both de-identified and publicly available and therefore we did not need to get IRB approval for this study.

## Data Availability Statement

Both cohorts used in our study are publicly available at no cost upon completion of the respective data-use agreements. We used all the T1 MRI scan and clinical data from ADNI (June 15th 2019 freeze), and NACC (Jan 5th 2019 freeze). The IDs of patients and scans used in our study and training, validation, test indicators are available in our github repository. Results of our Freesurfer segmentation (results of over 60,000 compute time on our HPC) for all of NACC and ADNI scans are also publicly available at no cost through NACC and ADNI websites, to anyone who has signed ADNI and NACC data-use agreements. Our trained model, training and validation scripts, and model predictions for each scan ID and patient ID, as well as Freesurfer segmentation volume and thickness features are also available as open-source on github.

## Author Contribution

CFG and NR co-led and supervised this study over all steps from design, development, and analysis. SL performed the majority of the data preprocessing, all of deep learning and baseline machine learning model development, training, validation and analysis. AVM provided clinical supervision throughout the study from design to analysis. HR provided neuro-imaging supervision throughout the study from design to analysis. JC developed Freesurfer segmentation scripts, performed quality control analysis of the Freesurfer results, contributed to writing and analysis of the results overall, and provided neuro-imaging supervision during the study. BZ developed scalable Freesurfer segmentation scripts, performed all Freesurfer segmentation jobs in our institution’s high performance cluster. WZ performed pre-processing of all scans in the NACC dataset. All authors participated in the writing of the manuscript.

## Acknowledgements

SL was partially supported by NSF DMS-2009752, NSF NRT-HDR-1922658, and Leon Lowenstein Foundation. NR was partially supported by Leon Lowenstein Foundation and NIH/NIA P30AG066512. AVM was partially supported by NIH/NIA P30AG008051 and P30AG066512. HR was partially supported by NIH/NIA P30AG066512 and NIA/NIBIB U24EB028980. JC was partially supported by NIH/NIA P30AG066512 and NIH-NINDS 1RF1NS110041-01.

The NACC database is funded by NIA/NIH Grant U01 AG016976. NACC data are contributed by the NIA-funded ADRCs: P30 AG019610 (PI Eric Reiman, MD), P30 AG013846 (PI Neil Kowall, MD), P50 AG008702 (PI Scott Small, MD), P50 AG025688 (PI Allan Levey, MD, PhD), P50 AG047266 (PI Todd Golde, MD, PhD), P30 AG010133 (PI Andrew Saykin, PsyD), P50 AG005146 (PI Marilyn Albert, PhD), P50 AG005134 (PI Bradley Hyman, MD, PhD), P50 AG016574 (PI Ronald Petersen, MD, PhD), P50 AG005138 (PI Mary Sano, PhD), P30 AG008051 (PI Thomas Wisniewski, MD), P30 AG013854 (PI Robert Vassar, PhD), P30 AG008017 (PI Jeffrey Kaye, MD), P30 AG010161 (PI David Bennett, MD), P50 AG047366 (PI Victor Henderson, MD, MS), P30 AG010129 (PI Charles DeCarli, MD), P50 AG016573 (PI Frank LaFerla, PhD), P50 AG005131 (PI James Brewer, MD, PhD), P50 AG023501 (PI Bruce Miller, MD), P30 AG035982 (PI Russell Swerdlow, MD), P30 AG028383 (PI Linda Van Eldik, PhD), P30 AG053760 (PI Henry Paulson, MD, PhD), P30 AG010124 (PI John Trojanowski, MD, PhD), P50 AG005133 (PI Oscar Lopez, MD), P50 AG005142 (PI Helena Chui, MD), P30 AG012300 (PI Roger Rosenberg, MD), P30 AG049638 (PI Suzanne Craft, PhD), P50 AG005136 (PI Thomas Grabowski, MD), P50 AG033514 (PI Sanjay Asthana, MD, FRCP), P50 AG005681 (PI John Morris, MD), P50 AG047270 (PI Stephen Strittmatter, MD, PhD).

Data collection and sharing for the ADNI part of this project was funded by the Alzheimer’s Disease Neuroimaging Initiative (ADNI) (National Institutes of Health Grant U01 AG024904) and DOD ADNI (Department of Defense award number W81XWH-12-2-0012). ADNI is funded by the National Institute on Aging, the National Institute of Biomedical Imaging and Bioengineering, and through generous contributions from the following: AbbVie, Alzheimer’s Association; Alzheimer’s Drug Discovery Foundation; Araclon Biotech; BioClinica, Inc.; Biogen; Bristol-Myers Squibb Company; CereSpir, Inc.; Cogstate; Eisai Inc.; Elan Pharmaceuticals, Inc.; Eli Lilly and Company; EuroImmun; F. Hoffmann-La Roche Ltd and its affiliated company Genentech, Inc.; Fujirebio; GE Healthcare; IXICO Ltd.;Janssen Alzheimer Immunotherapy Research & Development, LLC.; Johnson & Johnson Pharmaceutical Research & Development LLC.; Lumosity; Lundbeck; Merck & Co., Inc.;Meso Scale Diagnostics, LLC.; NeuroRx Research; Neurotrack Technologies; Novartis Pharmaceuticals Corporation; Pfizer Inc.; Piramal Imaging; Servier; Takeda Pharmaceutical Company; and Transition Therapeutics. The Canadian Institutes of Health Research is providing funds to support ADNI clinical sites in Canada. Private sector contributions are facilitated by the Foundation for the National Institutes of Health (www.fnih.org). The grantee organization is the Northern California Institute for Research and Education, and the study is coordinated by the Alzheimer’s Therapeutic Research Institute at the University of Southern California. ADNI data are disseminated by the Laboratory for Neuro Imaging at the University of Southern California.

## Footnotes

2 https://adni.loni.usc.edu/

3 https://developer.nvidia.com/cuda-toolkit

