## Supplementary material for "Development of a Deep Learning Model for Early Alzheimer’s Disease Detection from Structural MRIs and External Validation on an Independent Cohort": Alzheimer's paper supplementary.docx

Supplementary Table S1: Diagnosis Criteria for Cognitively Normal, Mild Cognitive Impairment and Mild Alzheimer’s Disease Dementia in two cohorts: ADNI and NACC.

| Criteria | ADNI^[1]^ | NACC^[2]^ |
| --- | --- | --- |
| Cognitively Normal | - No Memory Complaints aside from those common to other normal subjects of that age range. - Normal memory function documented by scoring at specific cutoffs on the Logical Memory II subscale (delayed Paragraph Recall) from the Wechsler Memory Scale - Revised (the maximum score is 25): a) greater than or equal to 9 for 16 or more years of education b) greater than or equal to 5 for 8-15 years of education c) greater than or equal to 3 for 0-7 years of education. - Mini-Mental State Exam score between 24 and 30 (inclusive) (Exceptions may be made for subjects with less than 8 years of education at the discretion of the project director). - Clinical Dementia Rating = 0. Memory Box score must be 0. - Cognitively normal, based on an absence of significant impairment in cognitive functions or activities of daily living. | The subject has normal cognition (global CDR=0 and/or neuropsychological testing within normal range) and normal behavior (i.e., the subject does not exhibit behavior sufficient to diagnose MCI or dementia due to FTLD or LBD) |
| MCI | - Memory complaint by subject or study partner that is verified by a study partner. - Abnormal memory function documented by scoring below the education adjusted cutoff on the Logical Memory II subscale (Delayed Paragraph Recall) from the Wechsler Memory Scale – Revised (the maximum score is 25): a) less than or equal to 8 for 16 or more years of education b) less than or equal to 4 for 8-15 years of education c) less than or equal to 2 for 0-7 years of education. - Mini-Mental State Exam score between 24 and 30 (inclusive) (Exceptions may be made for subjects with less than 8 years of education at the discretion of the project director). - Clinical Dementia Rating = 0.5. Memory Box score must be at least 0.5. - General cognition and functional performance sufficiently preserved such that a diagnosis of Alzheimer’s disease cannot be made by the site physician at the time of the screening visit. | If the subject does not have normal cognition or behavior and is not clinically demented, indicate the type of cognitive impairment below.  MCI CORE CLINICAL CRITERIA • Is the subject, the co-participant, or a clinician concerned about a change in cognition compared to the subject’s previous level?  • Is there impairment in one or more cognitive domains (memory, language, executive function, attention, and visuospatial skills)?  • Is there largely preserved independence in functional abilities (no change from prior manner of functioning or uses minimal aids or assistance)? |
| AD | - Memory complaint by subject or study partner that is verified by a study partner. - Abnormal memory function documented by scoring below the education adjusted cutoff on the Logical Memory II subscale (Delayed Paragraph Recall) from the Wechsler Memory Scale – Revised (the maximum score is 25): a) less than or equal to 8 for 16 or more years of education b) less than or equal to 4 for 8-15 years of education c) less than or equal to 2 for 0-7 years of education. - MMSE between 20 and 26 (inclusive) (Exceptions may be made for subjects with less than 8 years of education at the discretion of the protocol PI). - Clinical Dementia Rating = 0.5, 1.0 ➪ NINCDS/ADRDA criteria for probable AD. | The subject has cognitive or behavioral (neuropsychiatric) symptoms that meet all of the following criteria:  • Interfere with ability to function as before at work or at usual activities? Represent a decline from previous levels of functioning?  • Are not explained by delirium or major psychiatric disorder?  • Include cognitive impairment detected and diagnosed through a combination of 1) history-taking and 2) objective cognitive assessment (bedside or neuropsychological testing)?  AND Impairment in one* or more of the following domains.  – Impaired ability to acquire and remember new information  – Impaired reasoning and handling of complex tasks, poor judgment  – Impaired visuospatial abilities  – Impaired language functions  – Changes in personality, behavior, or comportment  * In the event of single-domain impairment (e.g., language in PPA, behavior in bvFTD, posterior cortical atrophy), the subject must not fulfill criteria for MCI. |

Supplementary Table S2: List of 138 volumes and thickness of clinically-relevant brain ROIs, obtained by Freesurfer for each included subject.

| **Volumes or Thickness Variables Based on Brain ROIs** | | | |
| --- | --- | --- | --- |
| Left-Lateral-Ventricle | Right-WM-hypointensities | lh_entorhinal_volume | rh_cuneus_volume |
| Left-Inf-Lat-Vent | non-WM-hypointensities | lh_fusiform_volume | rh_entorhinal_volume |
| Left-Cerebellum-White-Matter | Left-non-WM-hypointensities | lh_inferiorparietal_volume | rh_fusiform_volume |
| Left-Cerebellum-Cortex | Right-non-WM-hypointensities | lh_inferiortemporal_volume | rh_inferiorparietal_volume |
| Left-Thalamus-Proper | Optic-Chiasm | lh_isthmuscingulate_volume | rh_inferiortemporal_volume |
| Left-Caudate | CC_Posterior | lh_lateraloccipital_volume | rh_isthmuscingulate_volume |
| Left-Putamen | CC_Mid_Posterior | lh_lateralorbitofrontal_volume | rh_lateraloccipital_volume |
| Left-Pallidum | CC_Central | lh_lingual_volume | rh_lateralorbitofrontal_volume |
| 3rd-Ventricle | CC_Mid_Anterior | lh_medialorbitofrontal_volume | rh_lingual_volume |
| 4th-Ventricle | CC_Anterior | lh_middletemporal_volume | rh_medialorbitofrontal_volume |
| Brain-Stem | BrainSegVol | lh_parahippocampal_volume | rh_middletemporal_volume |
| Left-Hippocampus | BrainSegVolNotVent | lh_paracentral_volume | rh_parahippocampal_volume |
| Left-Amygdala | BrainSegVolNotVentSurf | lh_parsopercularis_volume | rh_paracentral_volume |
| CSF | lhCortexVol | lh_parsorbitalis_volume | rh_parsopercularis_volume |
| Left-Accumbens-area | rhCortexVol | lh_parstriangularis_volume | rh_parsorbitalis_volume |
| Left-VentralDC | CortexVol | lh_pericalcarine_volume | rh_parstriangularis_volume |
| Left-vessel | lhCerebralWhiteMatterVol | lh_postcentral_volume | rh_pericalcarine_volume |
| Left-choroid-plexus | rhCerebralWhiteMatterVol | lh_posteriorcingulate_volume | rh_postcentral_volume |
| Right-Lateral-Ventricle | CerebralWhiteMatterVol | lh_precentral_volume | rh_posteriorcingulate_volume |
| Right-Inf-Lat-Vent | SubCortGrayVol | lh_precuneus_volume | rh_precentral_volume |
| Right-Cerebellum-White-Matter | TotalGrayVol | lh_rostralanteriorcingulate_volume | rh_precuneus_volume |
| Right-Cerebellum-Cortex | SupraTentorialVol | lh_rostralmiddlefrontal_volume | rh_rostralanteriorcingulate_volume |
| Right-Thalamus-Proper | SupraTentorialVolNotVent | lh_superiorfrontal_volume | rh_rostralmiddlefrontal_volume |
| Right-Caudate | SupraTentorialVolNotVentVox | lh_superiorparietal_volume | rh_superiorfrontal_volume |
| Right-Putamen | MaskVol | lh_superiortemporal_volume | rh_superiorparietal_volume |
| Right-Pallidum | BrainSegVol-to-eTIV | lh_supramarginal_volume | rh_superiortemporal_volume |
| Right-Hippocampus | MaskVol-to-eTIV | lh_frontalpole_volume | rh_supramarginal_volume |
| Right-Amygdala | lhSurfaceHoles | lh_temporalpole_volume | rh_frontalpole_volume |
| Right-Accumbens-area | rhSurfaceHoles | lh_transversetemporal_volume | rh_temporalpole_volume |
| Right-VentralDC | SurfaceHoles | lh_insula_volume | rh_transversetemporal_volume |
| Right-vessel | EstimatedTotalIntraCranialVol | BrainSegVolNotVent | rh_insula_volume |
| Right-choroid-plexus | lh_bankssts_volume | eTIV |  |
| 5th-Ventricle | lh_caudalanteriorcingulate_volume | rh_bankssts_volume |  |
| WM-hypointensities | lh_caudalmiddlefrontal_volume | rh_caudalanteriorcingulate_volume |  |
| Left-WM-hypointensities | lh_cuneus_volume | rh_caudalmiddlefrontal_volume |  |

Supplementary Table S3: Performance of deep learning model and the ROI-volume/thickness model on patient subgroups based on Gender, Education and ApoE4 status.

|  | ADNI Heldout Patients  (n=90 individuals, 297 scans) | | NACC external validation  (n=1522 individuals, 2025 scans ) | |
| --- | --- | --- | --- | --- |
|  | Deep learning model  Area under ROC curve | ROI-volume/thickness  Area under ROC curve | Deep learning model  Area under ROC curve | ROI-volume/thickness  Area under ROC curve |
| **Gender: Male** |  |  |  |  |
| Cognitively Normal | 88.42  (95% CI: 87.51-89.34) | 84.95  (95% CI: 84.11-85.79) | 85.82  (95% CI: 85.50-86.14) | 82.97  (95% CI: 82.48-83.47) |
| Mild Cognitive Impairment | 57.59  (95% CI: 55.89-59.30) | 55.19  (95% CI: 53.71-56.66) | 57.81  (95% CI: 57.29-58.33) | 57.94  (95% CI: 57.40-58.47) |
| Alcheimer’s Disease Dementia | 90.62  (95% CI: 89.64-91.61) | 77.79  (95% CI: 77.04-78.55) | 90.84  (95% CI: 90.68-91.00) | 82.53  (95% CI: 82.01-83.06) |
| **Gender: Female** |  |  |  |  |
| Cognitively Normal | 92.85  (95% CI: 92.18-93.53) | 83.96  (95% CI: 83.21-84.72) | 85.15  (95% CI: 84.67-85.63) | 79.88  (95% CI: 79.54-80.23) |
| Mild Cognitive Impairment | 66.35  (95% CI: 64.53-68.16) | 63.85  (95% CI: 62.44-65.27) | 63.20  (95% CI: 62.92-63.48) | 57.54  (95% CI: 56.97-58.11) |
| Alcheimer’s Disease Dementia | 99.21  (95% CI: 98.70-99.71) | 89.90  (95% CI: 89.10-90.70) | 92.89  (95% CI: 92.46-93.33) | 81.79  (95% CI: 81.30-82.27) |
| **Education <15 Years** |  |  |  |  |
| Cognitively Normal | 94.86  (95% CI: 94.39-95.34) | 93.34  (95% CI: 92.58-94.10) | 86.19  (95% CI: 85.77-86.62) | 79.62  (95% CI: 79.15-80.10) |
| Mild Cognitive Impairment | 52.88  (95% CI: 50.74-55.02) | 66.70  (95% CI: 65.04-68.35) | 55.11  (95% CI: 54.42-55.80) | 60.22  (95% CI: 59.25-61.20) |
| Alcheimer’s Disease Dementia | 88.42  (95% CI: 87.38-89.46) | 86.90  (95% CI: 85.92-87.88) | 89.90  (95% CI: 89.54-90.26) | 80.59  (95% CI: 80.06-81.12) |
| **Education >=15 Years** |  |  |  |  |
| Cognitively Normal | 86.66  (95% CI: 85.75-87.58) | 82.25  (95% CI: 81.17-83.33) | 84.38  (95% CI: 84.18-84.58) | 81.90  (95% CI: 81.46-82.34) |
| Mild Cognitive Impairment | 63.64  (95% CI: 62.69-64.60) | 55.50  (95% CI: 54.04-56.95) | 62.58  (95% CI: 61.94-63.23) | 56.12  (95% CI: 55.55-56.69) |
| Alcheimer’s Disease Dementia | 92.73  (95% CI: 92.19-93.28) | 82.70  (95% CI: 82.02-83.38) | 92.90  (95% CI: 92.67-93.12) | 83.38  (95% CI: 83.02-83.73) |
| **With ApoE4** |  |  |  |  |
| Cognitively Normal | 63.82  (95% CI: 62.11-65.52) | 80.92  (95% CI: 79.80-82.05) | 83.57  (95% CI: 83.24-83.91) | 84.69  (95% CI: 84.32-85.06) |
| Mild Cognitive Impairment | 47.47  (95% CI: 46.61-48.32) | 64.19  (95% CI: 63.27-65.11) | 57.34  (95% CI: 56.79-57.90) | 58.37  (95% CI: 57.98-58.77) |
| Alcheimer’s Disease Dementia | 81.78  (95% CI: 81.03-82.54) | 75.16  (95% CI: 74.30-76.01) | 90.29  (95% CI: 90.03-90.54) | 84.70  (95% CI: 84.30-85.10) |
| **Without ApoE4** |  |  |  |  |
| Cognitively Normal | 82.37  (95% CI: 81.67-83.07) | 72.82  (95% CI: 72.31-73.33) | 87.32  (95% CI: 87.13-87.51) | 78.57  (95% CI: 78.43-78.71) |
| Mild Cognitive Impairment | 68.57  (95% CI: 67.59-69.56) | 52.88  (95% CI: 51.96-53.80) | 64.65  (95% CI: 64.27-65.03) | 57.56  (95% CI: 57.31-57.82) |
| Alcheimer’s Disease Dementia | 91.74  (95% CI: 90.82-92.65) | 81.18  (95% CI: 80.19-82.18) | 92.60  (95% CI: 92.42-92.77) | 79.90  (95% CI: 79.68-80.12) |

Supplementary Table S4: Importance of ROIs in deep learning and ROI-volume/thickness model in classification of CN/MCI/AD. Details of our approach to compute importances is in the methods section.

| **Rank** | **ROIs** | **CN**  **Normed Gradient Count** | **MCI**  **Normed Gradient Count** | **AD**  **Normed Gradient Count** | **ROI model Feature Impor.** | **Rank** | **ROIs** | **CN**  **Normalized Gradient Count** | **MCI**  **Normed Gradient Count** | **AD**  **Normed Gradient Count** | **ROI model Feature Import.** |
| --- | --- | --- | --- | --- | --- | --- | --- | --- | --- | --- | --- |
| 1 | **4th-Ventricle** | 12.55 | 8.23 | 13.23 | 0.02 | 52 | **rh_cuneus** | 7.97 | 2.25 | 7.18 | 0.00 |
| 2 | **Left-Hippocampus** | 9.93 | 2.33 | 11.86 | 0.12 | 53 | **Right-Accumbens-area** | 6.31 | 2.81 | 7.85 | 0.01 |
| 3 | **rh_parahippocampal** | 10.01 | 2.19 | 11.84 | 0.01 | 54 | **rh_lateralorbitofrontal** | 7.85 | 1.95 | 7.28 | 0.00 |
| 4 | **CSF** | 10.88 | 3.03 | 11.75 | 0.00 | 55 | **lh_temporalpole** | 7.67 | 0.81 | 7.84 | 0.01 |
| 5 | **rh_isthmuscingulate** | 10.22 | 4.04 | 11.67 | 0.01 | 56 | **rh_precentral** | 7.78 | 1.27 | 7.69 | 0.00 |
| 6 | **lh_parahippocampal** | 10.03 | 1.94 | 11.65 | 0.02 | 57 | **lh_medialorbitofrontal** | 7.60 | 1.87 | 7.30 | 0.00 |
| 7 | **rh_entorhinal** | 9.78 | 0.38 | 11.55 | 0.02 | 58 | **rh_superiortemporal** | 7.32 | 0.41 | 7.59 | 0.01 |
| 8 | **lh_entorhinal** | 10.12 | 0.60 | 11.54 | 0.02 | 59 | **Right-Pallidum** | 6.76 | 0.87 | 7.47 | 0.01 |
| 9 | **lh_transversetemporal** | 11.43 | 2.54 | 11.49 | 0.00 | 60 | **lh_pericalcarine** | 7.38 | 1.93 | 7.45 | 0.00 |
| 10 | **Right-Amygdala** | 9.03 | 2.99 | 11.40 | 0.01 | 61 | **Left-choroid-plexus** | 7.31 | 0.93 | 7.34 | 0.00 |
| 11 | **Right-Hippocampus** | 9.38 | 1.31 | 11.28 | 0.03 | 62 | **Right-Lateral-Ventricle** | 7.33 | 3.00 | 6.38 | 0.01 |
| 12 | **WM-hypointensities** | 11.12 | 3.22 | 9.48 | 0.03 | 63 | **rh_supramarginal** | 6.99 | 1.51 | 7.21 | 0.00 |
| 13 | **rh_transversetemporal** | 10.24 | 1.36 | 11.07 | 0.00 | 64 | **lh_postcentral** | 7.18 | 1.55 | 6.91 | 0.00 |
| 14 | **rh_posteriorcingulate** | 10.89 | 2.07 | 11.06 | 0.00 | 65 | **rh_postcentral** | 6.92 | 1.10 | 7.17 | 0.01 |
| 15 | **lh_isthmuscingulate** | 9.93 | 4.18 | 11.00 | 0.01 | 66 | **lh_cuneus** | 7.14 | 1.99 | 6.84 | 0.02 |
| 16 | **lh_insula** | 10.38 | 1.92 | 10.71 | 0.00 | 67 | **Left-Lateral-Ventricle** | 7.14 | 0.99 | 6.30 | 0.01 |
| 17 | **CC_Posterior** | 9.57 | 1.88 | 10.42 | 0.01 | 68 | **lh_rostralanteriorcingulate** | 7.03 | 2.10 | 5.77 | 0.01 |
| 18 | **Left-Amygdala** | 8.25 | 3.45 | 10.42 | 0.07 | 69 | **rh_caudalmiddlefrontal** | 6.97 | 0.81 | 6.54 | 0.00 |
| 19 | **lh_posteriorcingulate** | 9.59 | 2.66 | 10.30 | 0.00 | 70 | **lh_lateralorbitofrontal** | 6.91 | 1.42 | 6.08 | 0.00 |
| 20 | **rh_insula** | 9.70 | 1.51 | 10.29 | 0.00 | 71 | **Left-Pallidum** | 6.79 | 1.58 | 6.91 | 0.01 |
| 21 | **Right-VentralDC** | 9.30 | 1.35 | 10.07 | 0.00 | 72 | **lh_superiortemporal** | 6.90 | 0.58 | 6.79 | 0.01 |
| 22 | **rh_precuneus** | 9.73 | 2.89 | 10.05 | 0.02 | 73 | **lh_inferiortemporal** | 6.29 | 0.42 | 6.72 | 0.00 |
| 23 | **rh_caudalanteriorcingulate** | 9.80 | 0.82 | 9.08 | 0.01 | 74 | **lh_superiorfrontal** | 6.59 | 0.68 | 5.18 | 0.00 |
| 24 | **Right-Thalamus-Proper** | 9.01 | 1.78 | 9.76 | 0.00 | 75 | **rh_medialorbitofrontal** | 6.52 | 2.20 | 5.99 | 0.00 |
| 25 | **rh_bankssts** | 9.10 | 1.41 | 9.72 | 0.01 | 76 | **rh_superiorfrontal** | 6.41 | 0.56 | 5.30 | 0.01 |
| 26 | **Left-Caudate** | 9.40 | 2.14 | 9.67 | 0.00 | 77 | **Right-choroid-plexus** | 6.37 | 3.05 | 5.96 | 0.00 |
| 27 | **rh_lingual** | 9.12 | 2.65 | 9.54 | 0.00 | 78 | **lh_superiorparietal** | 6.35 | 0.75 | 5.57 | 0.01 |
| 28 | **Left-VentralDC** | 8.69 | 1.62 | 9.52 | 0.00 | 79 | **lh_supramarginal** | 6.18 | 1.49 | 6.32 | 0.01 |
| 29 | **CC_Mid_Posterior** | 9.50 | 1.78 | 9.21 | 0.00 | 80 | **rh_superiorparietal** | 6.19 | 1.20 | 5.57 | 0.01 |
| 30 | **lh_fusiform** | 8.42 | 1.28 | 9.47 | 0.01 | 81 | **rh_inferiortemporal** | 6.14 | 0.43 | 6.14 | 0.01 |
| 31 | **3rd-Ventricle** | 9.32 | 4.15 | 8.91 | 0.01 | 82 | **rh_middletemporal** | 6.05 | 0.51 | 6.12 | 0.02 |
| 32 | **Left-Thalamus-Proper** | 8.55 | 2.23 | 9.32 | 0.00 | 83 | **Left-Accumbens-area** | 5.11 | 1.45 | 6.02 | 0.00 |
| 33 | **lh_parsopercularis** | 9.30 | 2.19 | 8.53 | 0.01 | 84 | **rh_parstriangularis** | 5.93 | 0.58 | 5.50 | 0.01 |
| 34 | **rh_pericalcarine** | 9.29 | 3.14 | 9.26 | 0.00 | 85 | **Right-Cerebellum-Cortex** | 5.80 | 0.84 | 5.47 | 0.00 |
| 35 | **CC_Mid_Anterior** | 9.17 | 1.81 | 9.26 | 0.01 | 86 | **rh_inferiorparietal** | 5.58 | 1.71 | 5.72 | 0.00 |
| 36 | **Right-Caudate** | 9.20 | 2.45 | 9.04 | 0.00 | 87 | **lh_parstriangularis** | 5.69 | 0.91 | 4.00 | 0.02 |
| 37 | **rh_parsopercularis** | 9.03 | 1.52 | 9.15 | 0.00 | 88 | **Right-CerebellumWhitMtr** | 5.68 | 0.40 | 5.52 | 0.00 |
| 38 | **lh_precuneus** | 8.79 | 2.34 | 9.15 | 0.01 | 89 | **Left-Cerebellum-Cortex** | 5.38 | 0.90 | 5.57 | 0.00 |
| 39 | **lh_lingual** | 8.52 | 2.29 | 9.10 | 0.01 | 90 | **Left-Putamen** | 5.33 | 1.98 | 5.52 | 0.00 |
| 40 | **lh_caudalanteriorcingulate** | 8.99 | 1.27 | 8.47 | 0.01 | 91 | **lh_middletemporal** | 5.37 | 0.37 | 5.49 | 0.01 |
| 41 | **rh_paracentral** | 8.89 | 0.63 | 8.67 | 0.01 | 92 | **Left-CerebellumWhiteMTr** | 5.36 | 0.65 | 5.46 | 0.00 |
| 42 | **CC_Central** | 8.84 | 0.50 | 8.27 | 0.00 | 93 | **lh_inferiorparietal** | 5.13 | 0.83 | 5.30 | 0.01 |
| 43 | **CC_Anterior** | 8.77 | 4.72 | 8.41 | 0.01 | 94 | **Right-Putamen** | 4.96 | 1.14 | 5.27 | 0.00 |
| 44 | **lh_paracentral** | 8.44 | 0.96 | 8.47 | 0.01 | 95 | **rh_lateraloccipital** | 5.18 | 0.90 | 3.90 | 0.00 |
| 45 | **rh_rostralanteriorcingulate** | 8.45 | 2.28 | 6.99 | 0.02 | 96 | **rh_parsorbitalis** | 4.79 | 0.43 | 4.08 | 0.01 |
| 46 | **rh_fusiform** | 8.08 | 1.23 | 8.39 | 0.01 | 97 | **lh_rostralmiddlefrontal** | 4.68 | 0.46 | 3.30 | 0.00 |
| 47 | **lh_bankssts** | 8.31 | 2.08 | 8.02 | 0.01 | 98 | **lh_lateraloccipital** | 4.23 | 0.29 | 3.76 | 0.02 |
| 48 | **lh_caudalmiddlefrontal** | 8.29 | 1.20 | 6.78 | 0.01 | 99 | **bk/others** | 4.23 | 0.67 | 3.96 |  |
| 49 | **rh_temporalpole** | 7.92 | 0.87 | 8.24 | 0.00 | 100 | **rh_rostralmiddlefrontal** | 3.97 | 0.20 | 3.79 | 0.00 |
| 50 | **Brain-Stem** | 7.73 | 0.88 | 8.10 | 0.01 | 101 | **lh_parsorbitalis** | 3.57 | 0.11 | 2.84 | 0.00 |
| 51 | **lh_precentral** | 8.06 | 1.30 | 7.39 | 0.00 |  |  |  |  |  |  |

Supplementary Table S5: Confusion matrix of the deep learning model in classification of CN/MCI/AD on ADNI heldout set. Rows represent the diagnosed classes and columns represent predictions of the model.

|  | **CN** | **MCI** | **AD** |
| --- | --- | --- | --- |
| **CN** | 76 | 9 | 4 |
| **MCI** | 34 | 44 | 33 |
| **AD** | 4 | 7 | 86 |

Supplementary Table S6: Confusion matrix of the deep learning model in classification of CN/MCI/AD on NACC dataset. Rows represent the diagnosed classes and columns represent predictions of the model.

|  | **CN** | **MCI** | **AD** |
| --- | --- | --- | --- |
| **CN** | 1147 | 88 | 46 |
| **MCI** | 164 | 55 | 103 |
| **AD** | 78 | 52 | 312 |

**ADNI	NACC**

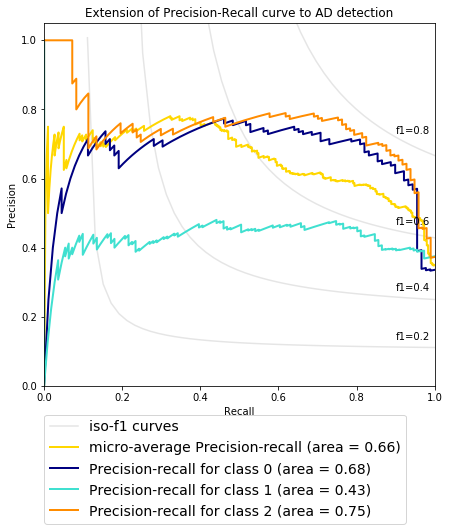

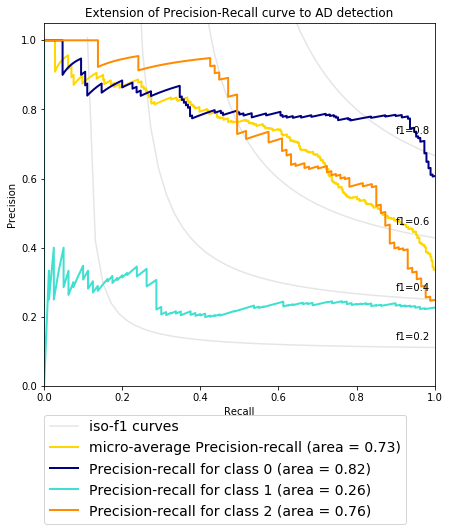

Supplementary Figure F1: Precision Recall Curve for prediction of CN (Class 0), MCI (Class 1) and AD (Class 2) in ADNI heldout test set and NACC external validation cohorts.

1. Reference: <http://adni.loni.usc.edu/wp-content/uploads/2010/09/ADNI_GeneralProceduresManual.pdf>
2. Reference: <https://files.alz.washington.edu/documentation/uds3-ivp-packet.pdf> and <https://r2d2.kumc.edu/ADC/Protocols/UDS2_protocol/ResearchDataDict_2015Mar.pdf> (column: NACCUDSD)
